# Prevalence and Severity of Anxiety, Depression, and Stress Among Optometry Students in Nigeria: A Cross-Sectional Study

**DOI:** 10.1101/2025.10.23.25338666

**Authors:** Michael Agyemang Kwarteng, Osamudiamen McHillary Ogiemudia, Bernadine N. Ekpenyong, Okechi U. Amaechi, Grace Ogbonna, Edith I. Daniel-Nwosu, Ngozika Esther Ezinne, Oforbuike Onyebuchi Ike, Kelechi C. Ogbuechi, Uchechukwu Levi Osuagwu

## Abstract

**Purpose:** To assess the prevalence and severity of anxiety, depression, and stress among optometry students in Nigeria, and to identify demographic factors associated with mental distress.

**Methods:** A cross-sectional, web-based survey was administered to optometry students from 11 Nigerian universities between 16 April and 18 November 2024. Mental health status was evaluated using the Depression, Anxiety, and Stress Scale-21 (DASS-21). Collected data included age, gender, year of study, marital status, and DASS-21 scores. Descriptive statistics summarized demographic characteristics and the prevalence of symptoms. Binary logistic regression was used to identify associations between demographic variables and mental health outcomes, with significance set at p<0.05.

**Results:** A total of 474 students participated (mean age: 23±3 years; 54.6% female; 94.1% unmarried). Overall, 51.1% experienced major depression, with 14.6% reporting severe or extremely severe symptoms. Severe anxiety was reported by 36.5%, and 25.1% experienced concurrent symptoms of anxiety, depression, and stress. Nearly half (49.2%) had at least two coexisting mental health conditions, indicating a significant emotional burden and potential impact on academic performance. Female students had significantly higher odds of experiencing depression (OR: 1.66; 95% CI:1.15–2.39) and overall mental distress (OR: 1.58; 95% CI: 1.03–2.43) compared to males.

**Conclusion:** This study revealed a high prevalence of mental health conditions among Nigerian optometry students, with one in four experiencing a comorbid of all three conditions. Female students were more likely to report adverse mental health outcomes. These findings underscore the urgent need for structured mental health support and preventive interventions within optometry training programs in Nigeria.

## 1. Introduction

The global burden of mental health conditions such as depression, anxiety, and stress continues to rise, significantly affecting the quality of life, academic productivity, and long-term health outcomes of sufferers (1). Mental health is increasingly being understood not only as the absence of psychological disorders but also as a state of well-being wherein individuals realize their abilities, can cope with everyday stresses, work productively, and contribute to their communities (2). Recent estimates suggest that one in every eight people worldwide lives with a mental disorder, with depression and anxiety being the most common (3).

The prevalence of mental health disorders among the general population, including elementary and secondary school students, has gained considerable attention, with several studies conducted among the population. However, other than in the USA, fewer studies have been done among college students concerning their mental health (4,5). Nevertheless, there is growing evidence of heightened vulnerability to mental health challenges among university students, particularly those in high-demand academic programs like health sciences. Stressors such as academic overload, clinical demands, examination pressure, and future career uncertainties have been identified as precipitating factors for psychological distress among medical and allied health students (6–9). While optometry students are a key segment within health professional training, they have received relatively less attention in mental health research, especially in low- and middle-income countries like Nigeria.

With a population of over 200 million, Nigeria faces a growing demand for quality eye care service delivery, necessitating the training of more optometrists across the country. Under the regulations of the Optometrists and Dispensing Opticians Registration Board of Nigeria (ODORBN), the optometry program has expanded from 3 to 14 tertiary institutions in the last four decades, with over 3,000 students currently enrolled nationwide (10,11). The Nigerian Optometry curriculum is structured as a six-year Doctor of Optometry program combining both academic and clinical training, and research, followed by a mandatory 12-month internship (11,12).

Despite the rigorous nature of the optometry program in Nigeria, the institutions continue to boast high numbers of students’ enrolment; however, this is often marred by high student drop-out and failure rates. Yet, there remains a dearth of data regarding the mental health outcomes and overall well-being of students enrolled in Optometry programs across Nigeria. Studies from related disciplines suggest that female students, students in clinical years, and those with limited social or financial support are disproportionately affected by poor mental health outcomes (13,14). A global review by Dyrbye et al.(7) found that health professional students face an elevated risk for burnout, suicidal ideation, and psychological comorbidities— highlighting the urgency for timely interventions.

The study by Ebeigbe et al.(15), which revealed notably high prevalence rates of depression (40.2%), anxiety (51.2%), and stress (35.5%) among optometry students at the University of Benin, strongly highlights the urgent need for a comprehensive national investigation into the mental health status of optometry students across Nigeria. Their findings also demonstrated associations between poor mental health outcomes and socio-demographic variables such as academic performance, financial background, social support and family dynamics. These findings echo those of studies conducted in other academic disciplines within the University (12), further suggesting that the academic environment in Nigerian universities may be a significant contributor to student mental health challenges. Complementing these, a recent cross-national study by Ike et al. (16) on university students and staff across sub-Saharan Africa found that Nigerian respondents had among the highest reported levels of severe and extremely severe depression, anxiety, and stress, second only to Malawi. This underscores a systemic mental health burden in Nigerian higher education institutions. Together, these findings make a compelling case for a national study that not only quantifies the prevalence but also explores the contextual determinants of mental health among optometry students, to inform targeted interventions and institutional mental health policies.

Furthermore, the implications of poor mental health extend beyond academic failure to include increased risk of substance abuse, professional attrition, and long-term morbidity (Chan et al., 2023) Depression and anxiety are also known to have bidirectional relationships with educational attainment—where low academic achievement may predispose individuals to mental health challenges, and early-onset mental disorders may hinder educational progression (17,18). Gender also plays a critical role in the epidemiology of mental health. Studies consistently report that female students are more likely to report symptoms of depression and anxiety than males (8,19). The reasons are multifactorial and may include biological susceptibility, gender-based social stressors, and systemic inequalities in educational or economic access (20).

In addition to academic pressures, Nigerian optometry students face unique mental health challenges arising from socio-political instability, financial hardship, and limited access to mental health care. Frequent strikes by academic unions, most notably the Academic Staff Union of Universities (ASUU), often lead to prolonged university closures, disrupted academic calendars, and delays in program completion (21,22). These disruptions are frequently attributed to unresolved funding disputes, unfulfilled government agreements, and deteriorating infrastructure, factors that contribute significantly to student anxiety, depression, and academic burnout.

Other aggravating conditions include insecurity on campuses, limited availability of mental health services, and stigma surrounding help-seeking behaviour (23). Many students face social isolation due to fear of judgment, while others resort to maladaptive coping strategies such as substance use. These patterns are compounded by a general neglect of student mental health within institutional policies and support systems. Moreover, the COVID-19 pandemic further intensified these challenges, introducing new stressors such as online learning fatigue, social disconnection, and health-related anxiety (24,25). Optometry students, like their medical students’ counterparts, tend to be disproportionately affected due to the interruption of clinical training, which is essential to their professional development (26).

The present study is therefore timely and relevant. It seeks to provide evidence-based insights into the prevalence and severity of mental health conditions among optometry students in Nigeria, using the standardized assessment tool (DASS-21). By evaluating the socio-demographic determinants, this study will also contribute to the development of targeted mental health policies and wellness support strategies in Nigerian optometry schools. It aims to address an urgent gap in the literature while aligning with the goals of Universal Health Coverage (UHC) and the WHO’s Comprehensive Mental Health Action Plan 2013–2030, which calls for expanded research and services in youth mental health, particularly in low-and middle-income settings (2).

## 2. Subjects and Methods

### 2.1 Participants

This study focused exclusively on optometry students enrolled in optometry schools in Nigeria. Participants were recruited from institutions offering optometry programs, and the survey targeted individuals currently pursuing undergraduate optometry degrees in Nigeria.

### 2.2 Setting

Nigeria, the most populous country in Africa, had a gross domestic product (GDP) of $252.7 billion in 2022 and a per capita GDP of $1,100 (27). As a key Sub-Saharan African (SSA) country with a growing higher education sector, Nigeria provides a relevant setting for exploring the mental health status of optometry students. There are 14 optometry schools in Nigeria (10).

### 2.3 Study Design and Procedure

A web-based, cross-sectional survey was conducted using a convenience sampling approach from Tuesday, April 16th to Monday, November 18th, 2024. A validated, self-administered questionnaire was distributed in English via Google Forms. The survey link was shared through social media platforms (e.g., WhatsApp, Facebook) and institutional email networks, using a snowballing strategy to enhance reach.

All prospective participants received a brief online description of the study’s purpose and procedures before proceeding. Participation required electronic consent, and individuals had to respond affirmatively to continue to the questionnaire. The survey strictly followed the STROBE guidelines for cross-sectional studies (28).

### 2.4 Questionnaire

A validated, self-administered survey (29) was adapted to suit the study’s objectives. The first section collected sociodemographic data, including age, gender, university, year of study, and marital status.

#### 2.4.1 DASS-21 Survey Scale

The Depression, Anxiety, and Stress Scale-21 (DASS-21) is a well-established, psychometrically robust instrument used globally to measure the severity of these three negative emotional states (30). It enables researchers to examine mental health status with internal validity and cross-cultural adaptability (31). The DASS-21 scale has been validated in several developing country contexts and is appropriate for assessing student mental health in Nigeria (32). This instrument contains 21 items, divided equally across three subscales (depression, anxiety, stress), with responses on a 4-point Likert scale (0 = did not apply at all; 3 = applied very much). The classification thresholds were:

- Depression: 0–9 (normal), 10–13 (mild), 14–20 (moderate), 21–27 (severe), 28+ (extremely severe)
- Anxiety: 0–7 (normal), 8–9 (mild), 10–14 (moderate), 15–19 (severe), 20+ (extremely severe)
- Stress: 0–14 (normal), 15–18 (mild), 19–25 (moderate), 26–33 (severe), 34+ (extremely severe)

The questionnaire was pre-tested with 10 optometry students from a Nigerian university one week before the main study. Feedback was used to refine the tool. The Cronbach’s alpha coefficient for the DASS-21 in this cohort was 0.82, indicating good internal consistency.

### 2.5 Inclusion and Exclusion Criteria

Only responses from undergraduate optometry students currently enrolled in Nigerian universities were included. Participants had to provide informed consent to participate. Duplicate entries, identified through identical IP addresses and matching demographics, were filtered, and only the most complete entry was retained.

### 2.6 Sample Size Determination

The required sample size was calculated based on a 95% confidence level, 5% precision, and an estimated 50% prevalence of mental health symptoms (due to the absence of similar prior studies in this cohort)(33). After accounting for a 10% non-response (attrition) rate, the final calculated sample size was 461. A total of 474 valid responses were received and included in the final analysis.

#### 2.6.1 Reliability Test

The DASS-21 demonstrated strong reliability in this population. The overall Cronbach’s alpha was 0.942. The subscales for: Depression: α = 0.806, Anxiety: α = 0.873 and Stress: α = 0.871. Each subscale included 7 items.

### 2.7 Data Analysis

The DASS-21 scores were accessed within October 4th and November 18th, 2024 and were analysed as categorical variables. Mental health symptoms were dichotomized for analysis:

- Depression: 0–9 = No depression; 10+ = depression
- Anxiety: 0–7 = No anxiety; 8+ = Anxiety
- Stress: 0–14 = No stress; 15+ = Stress

Sociodemographic factors included age, gender, marital status, level of study, and school affiliation. Frequencies and percentages were used to summarize sample characteristics. Prevalence rates were estimated using binomial distributions with Clopper-Pearson confidence intervals. Bivariate analyses were used to examine associations between mental health outcomes and sociodemographic characteristics. Subsequently, multivariable logistic regression was conducted, adjusting for age, gender, marital status, level of study, and university. A p-value < 0.05 was considered statistically significant. Analyses were conducted using IBM SPSS v21.0 and R v4.0.3.

### 2.8 Ethical Considerations

Ethical approval was obtained from Nigerian institutional ethics boards, including Federal University of Technology Owerri, Nigeria (FUT/SOHT/REC/vol. 4/2), Abia State University, Uturu, Nigeria (ABSU/REC/OPT/002/2024), and Bayero University Kano, Nigeria (NHREC/BUK-HREC/476/10/2311). Verbal informed consent was obtained electronically by requiring participants to select “Yes” to the question: *“Do you consent to voluntarily participate in this survey?ȝ* The question was mandatory, ensuring participants could not proceed without consenting. Anonymised data were collected such that the authors had no access to information that could identify individual participants during or after data collection.

## 3. Results

A total of 474 participants from 11 universities were involved in this study (see Figure 1). Participants were mostly from Bayero University Kano (37.3%), Abia State University (25.3%), and Madonna University (14.1%).

**Figure 1:**
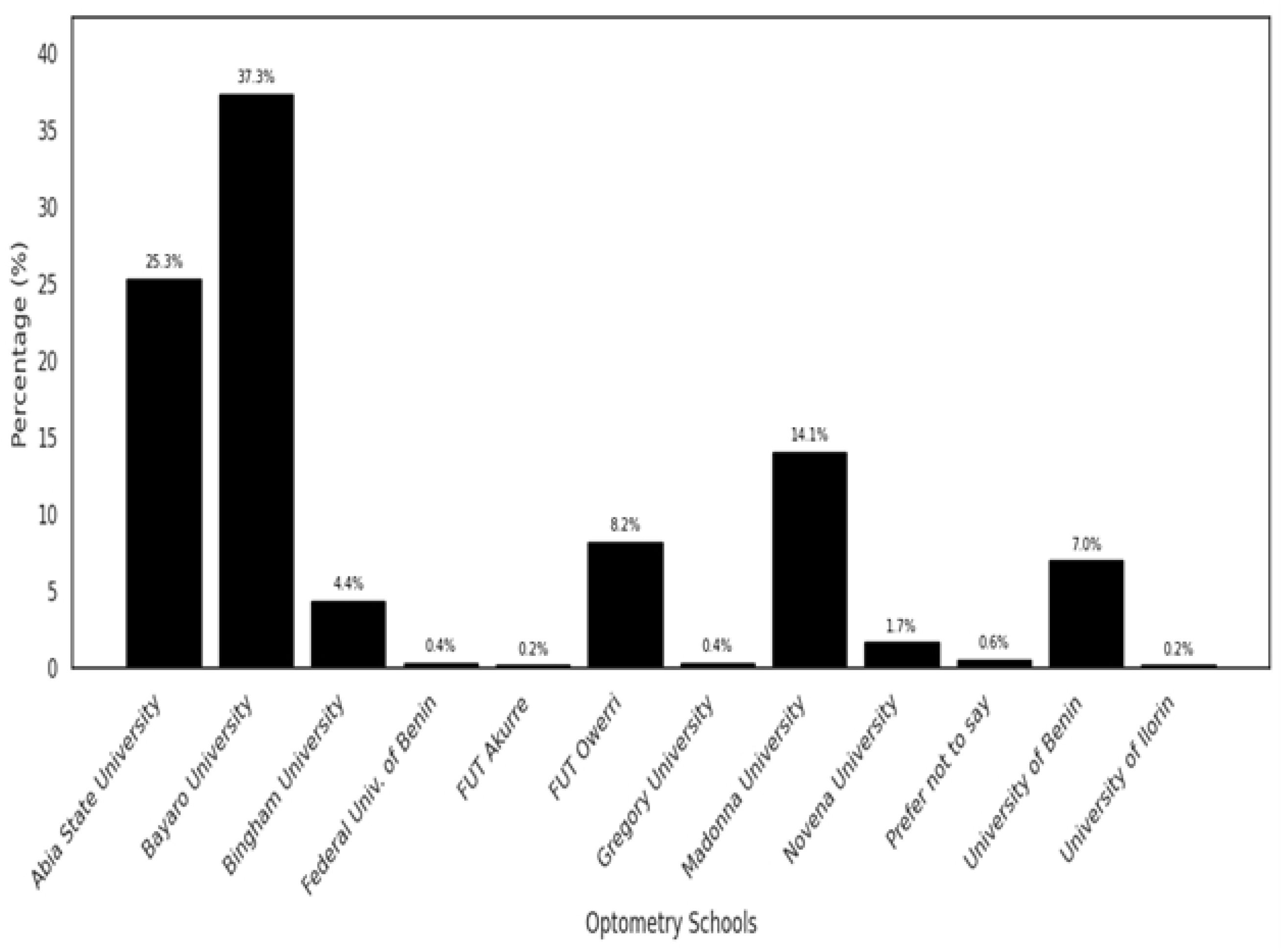
Distribution of participants by their institutions of learning. FUTO = Federal University of Technology.

The age of the participants ranged from 17 to 42 years (mean: 23 ± 3 years), mostly female (54.6%), not married (94.1%), and Christianity was the predominant religion (62.7%). In terms of year of study, 70.1% were in the clinical year, while 29.9% were in the preclinical year (years 1-4). Most respondents (95.1%) reported not having children, with only 4.9% indicating that they did, Table 1.

**Table 1:** Distribution of Demographic Characteristics.

| Variable | Category | Frequency (n) | Percentage (%) |
| --- | --- | --- | --- |
| Age group (years) | Younger age (<18yrs) | 5 | 1.1 |
|  | Youth (18-35yrs) | 467 | 98.5 |
| | Adult ($\geq 36$yrs) | 2 | 0.4 |
| Gender | Female | 259 | 54.6 |
|  | Male | 215 | 45.4 |
| Ethnicity | African descent | 467 | 98.5 |
|  | Others | 7 | 1.5 |
| Marital status | Married/relationship | 28 | 5.9 |
|  | Not married | 446 | 94.1 |
| Religion | Christianity | 297 | 62.7 |
|  | Islamic | 170 | 35.9 |
|  | Others | 7 | 1.5 |
| Year of study | Preclinical (Year 1-4) | 139 | 29.9 |
| (n=465) | Clinical (Year 5-6) | 326 | 70.1 |
| Have Children | Yes | 23 | 4.9 |
|  | No | 451 | 95.1 |

### 3.1 Distribution of the Severity of Mental Health Conditions

Figure 2 illustrates the severity distribution of anxiety, depression, and stress. Anxiety showed a notably high proportion of participants in the severe and extremely severe categories combined, totalling 36.5% (7.2% severe and 29.3% extremely severe), which is substantially higher compared to depression (14.6%) and stress (11.0%) in the same combined categories.

**Figure 2:**
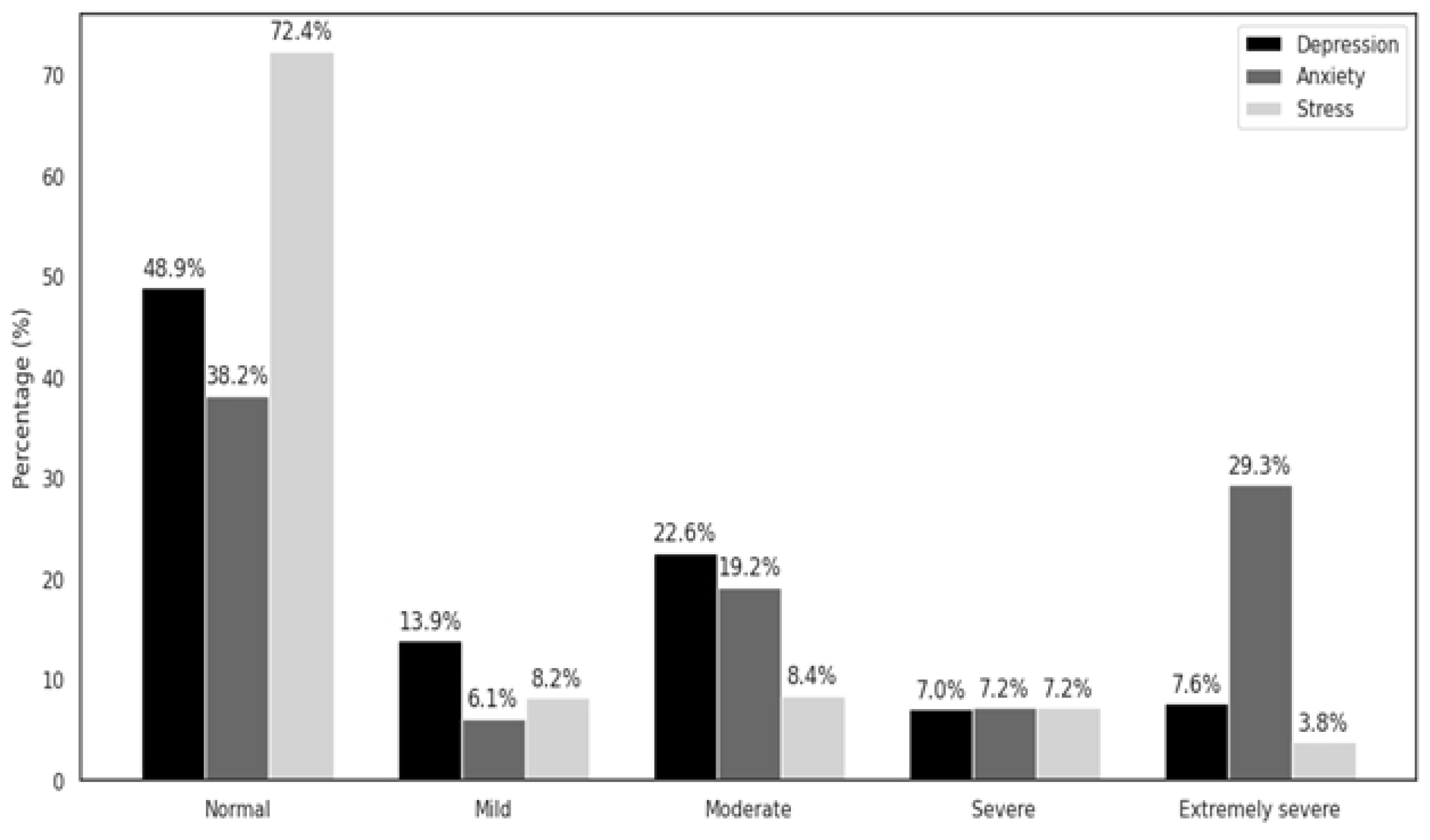
Distribution of the Severity of Mental Health Conditions

### 3.2 Distribution of the Prevalence of Mental Health Conditions

Over half of the participants reported experiencing a form of depression (51.1%) and anxiety (61.8%), while a smaller proportion reported stress (27.6%), Figure 3. Additionally, 25.1% of participants experienced all three conditions concurrently, and 49.2% experienced any two of these mental health issues. The prevalence of anxiety was notably higher compared to depression and stress, indicating a significant mental health burden.

**Figure 3:**
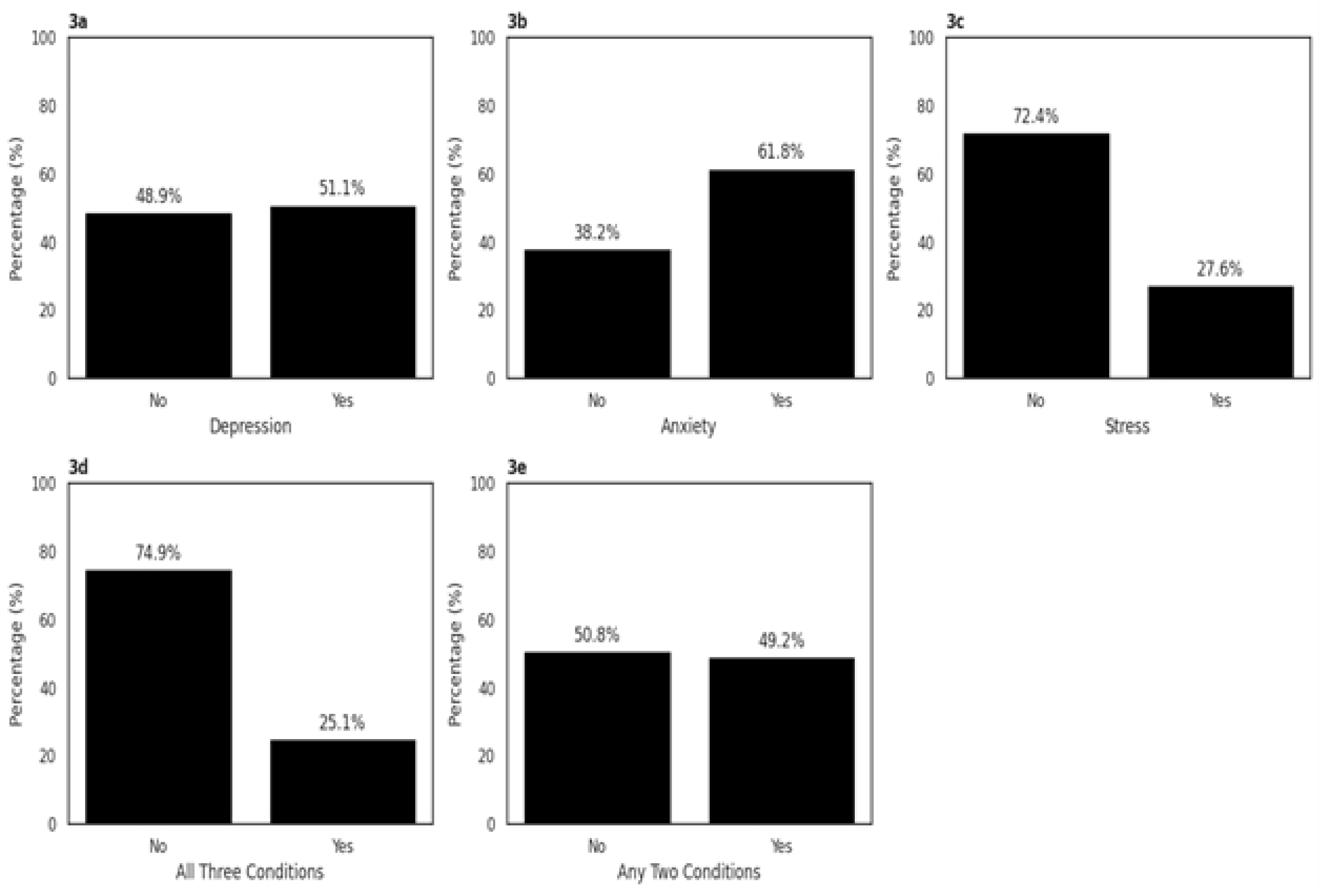
Distribution of the prevalence of Mental Health Conditions

### 3.3 Binary Logistic Regression Analysis of Mental Health Conditions and Demographics

A binary logistic regression was conducted to assess whether demographics predict mental health status. With each additional year of age, the odds of reporting symptoms of depression, anxiety, stress, or their co-occurrence decreased significantly, with odds ratios ranging from 0.91 to 0.93, Table 2. Female students had higher odds of depression (OR=1.66, 95%CI: 1.15 – 2.39) and experiencing all three mental health issues (OR=1.58, 95%CI: 1.03 – 2.43) compared to males. Being married was linked to lower odds of anxiety (OR=0.44, 95%CI: 0.20 – 0.95). Preclinical students, compared to clinical students, had higher odds of experiencing stress (OR = 1.59, 95% CI: 1.04–2.45), any two mental health issues (OR = 1.55, 95% CI: 1.04–2.31), and all three issues combined (OR = 1.61, 95% CI: 1.03–2.50), Table 2.

**Table 2:**
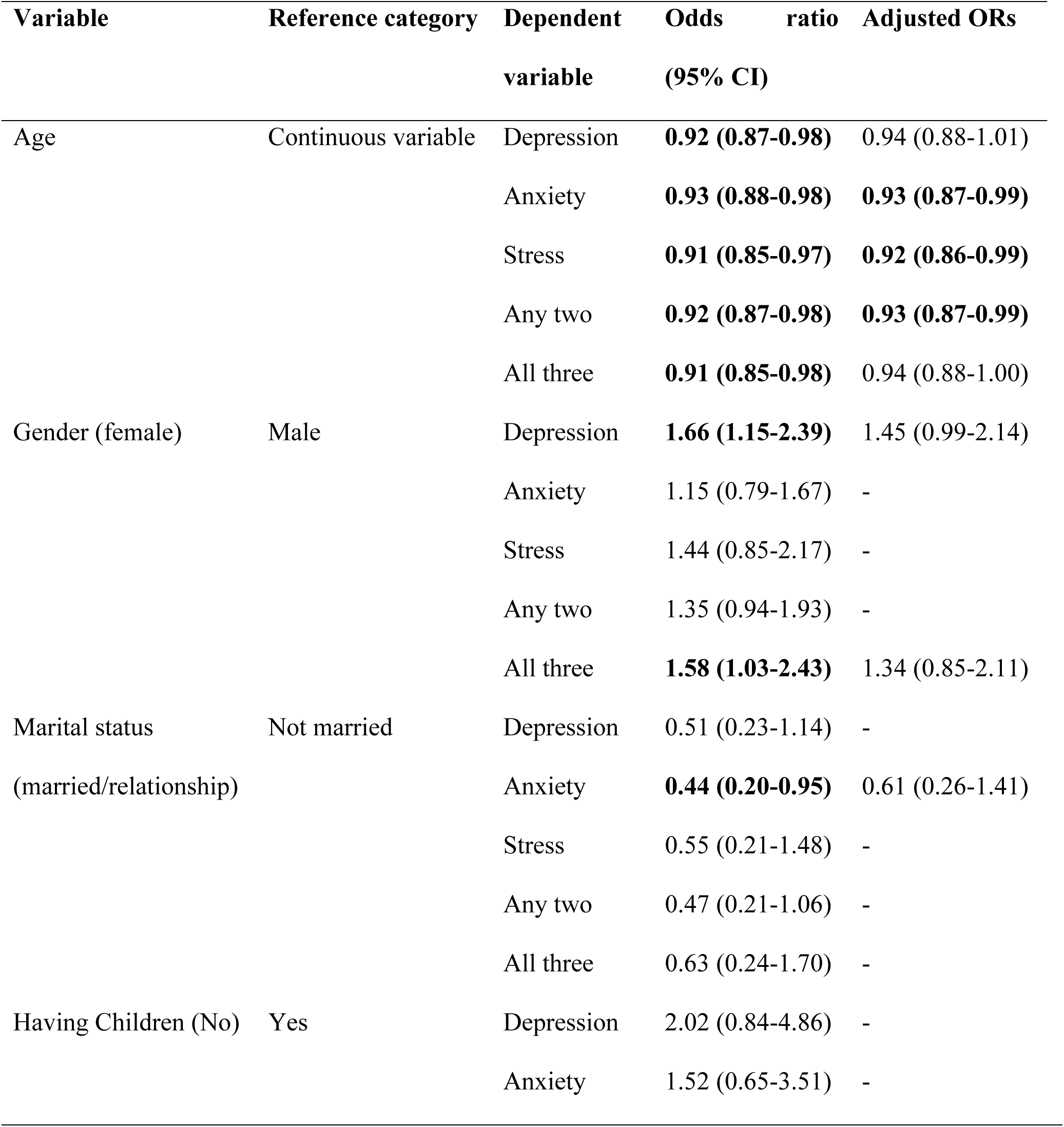
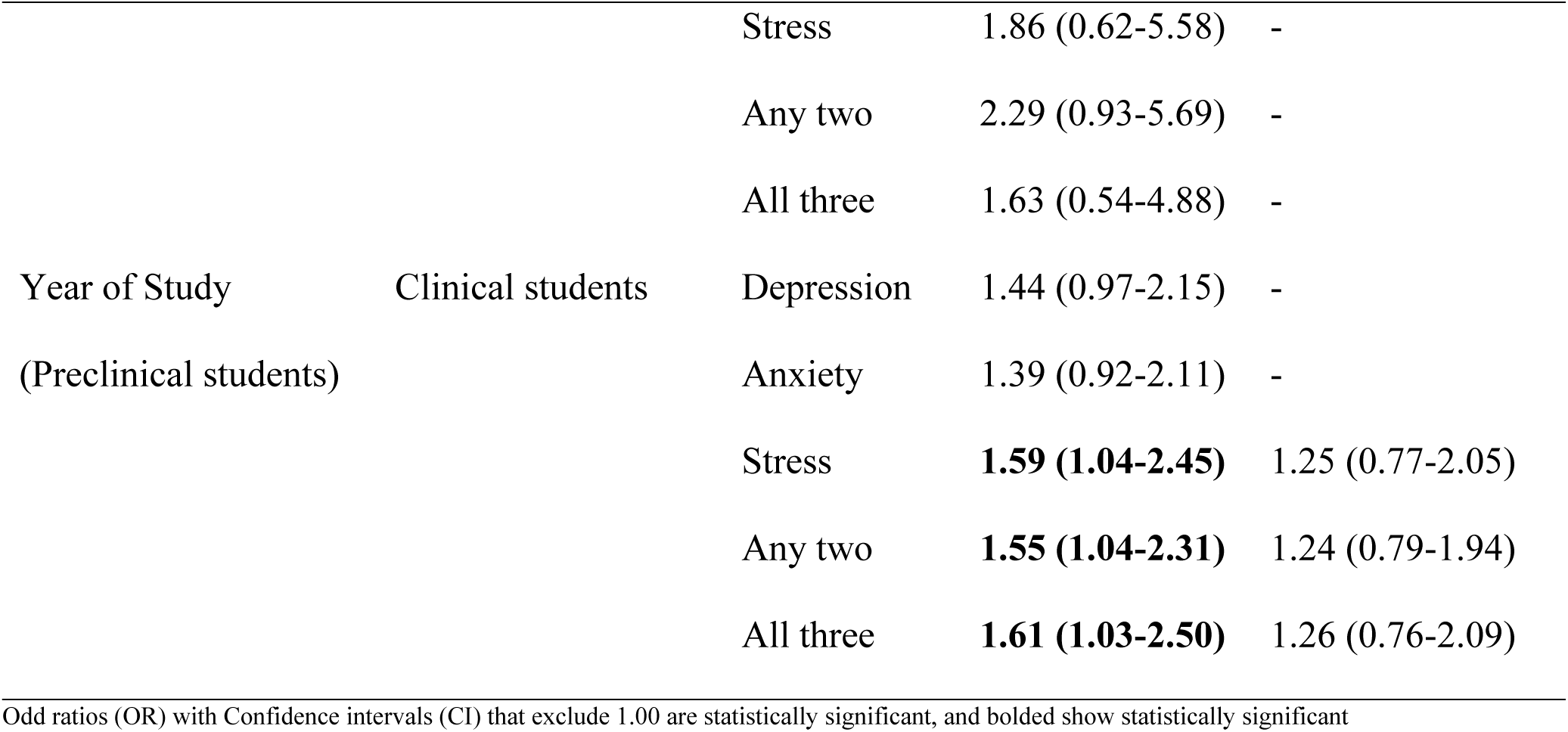
Binary Logistic Regression Analysis of Mental Health Conditions and Demographics.

After adjusting for significant variables such as age, year of study and marital status, the association between gender and depression attenuated, with the odds of depression among female students decreasing from an unadjusted OR of 1.66 (95% CI: 1.15–2.39) to an adjusted OR of 1.45 (95% CI: 0.99–2.14), rendering the association statistically non-significant. Similarly, the odds of experiencing all three mental health conditions among females declined from 1.58 (95% CI: 1.03–2.43) to AOR of 1.34 (95% CI: 0.85–2.11). Marital status was no longer a significant predictor of anxiety, with an adjusted OR of 0.61 (95% CI: 0.26–1.41). For year of study, clinical students initially showed higher odds of stress (OR = 1.59), any two conditions (OR = 1.55), and all three conditions (OR = 1.61); however, after adjusting for age, these associations weakened and were no longer significant (AORs = 1.25, 1.24, and 1.26, respectively).

## 4. Discussion

A significant mental health burden among this population was highlighted by examining the prevalence of anxiety, depression, and stress among university students in this study. The findings indicate a high prevalence of mental health challenges, especially anxiety, and emphasize key demographic factors influencing mental health outcomes. Anxiety emerged as the most common and severe mental health issue among participants, with a notable portion (61.8%) reporting symptoms and over one-third (36.5%) experiencing them at severe to extremely severe levels. This high prevalence highlights anxiety as a major mental health concern within the study population. These results align with previous studies (34–37), which also noted considerable variation in the severity of mental health symptoms, with anxiety often reported as the most prominent. Furthermore, to illustrate the scope of the problem, it has been reported that higher levels of depression, anxiety, and stress are closely linked to both suicidal ideation and academic underperformance in university students (38,39).

The study also showed the co-occurrence of mental health conditions in the population. A quarter of participants (25.1%) experienced anxiety, depression, and stress concurrently, and nearly half (49.2%) experienced any two conditions. This clustering of mental health conditions is consistent with the finding (40), which indicates a strong positive correlation among the three conditions, where an increase in one symptom is closely associated with significant increases in the others.

The significant demographic predictors of mental health outcomes in the current study were age, gender, and marital status. This study identified age as a significant protective factor against mental health challenges, with older students consistently showing lower odds of experiencing depression, anxiety, stress, or their co-occurrence (Table 2). Regarding gender, female students had significantly higher odds of experiencing depression and all three conditions concurrently. This finding aligns with previous research but differs from the study in which male students showed significantly higher levels of depression, anxiety, and stress. The study also highlighted the influence of marital status on mental health outcomes among university students. Consistent with previous research, results from the current study showed that married students are less likely to experience anxiety compared with unmarried students. This may reflect marital closeness, which provides some emotional and psychological stability, safety, and protection. However, it does not align with a report of a study (47) that showed higher levels of stress among married female students due to role conflicts in marriage. Nonetheless, the critical role of gender and societal expectations in a complex society like Nigeria cannot be overlooked. Unmarried students, especially females who are more vulnerable in academic settings, face intense pressure to marry by a certain age, increasing their likelihood of experiencing anxiety and/or academic stress. Additionally, they often assume caregiving responsibilities, which can further impact their mental well-being.

Regarding the year of study, findings revealed that students in their preclinical years were 1.6 times more likely to experience stress, comorbidity of any two conditions, and the presence of all three mental health conditions increased significantly than those in the clinical years. This may be due to preclinical students facing greater adjustment challenges, heavier theoretical coursework, and unfamiliar academic demands during the early years of training, which can increase stress levels and the risk of multiple mental health difficulties compared to students who have progressed to clinical years and may have developed better coping strategies and support networks. The findings of this study is consistent with other studies in which students in pre-clinical years had higher depression, anxiety, and stress scores than those in clinical years, but this differs from that observed in another research (52) that reported contrasting findings. Also, another study found that the year of study had no significant effect on outcome measures in mental health (53).

### 4.1 Strengths and Limitations

The study focused on the levels of mental health conditions among optometry students in Nigeria. The homogeneity within the sample supports more controlled analyses of variable factors such as marital status and year of study. The validated DASS-21 scale ensured that the data were reliable, and the web-based survey enabled widespread reach. However, the findings will be limited in their generalizability to the broader population of Nigerian students because of the use of the convenience sampling method. Optometry students in Nigeria largely belong to the relatively younger population, and this may affect how they experience and report mental health issues. The cross-sectional design limits the establishment of the cause-and-effect relationship of factors. Furthermore, the use of self-reported data introduces potential bias, especially given the significant stigma surrounding mental health in Nigerian society. Future research could benefit from incorporating qualitative data, such as responses to open-ended survey questions, to gain richer insights into respondents’ mental health-related beliefs and real-life experiences.

### 4.2 Conclusions

This study revealed a substantial burden of mental health conditions among Nigerian optometry students, with anxiety being the most prevalent. A significant proportion of students experience multiple co-occurring conditions, with one in four reporting concurrent anxiety, depression, and stress. Female students and those in preclinical years were identified as high-risk groups, while being older or married appeared to be protective. The findings underscore the need for integrated institutional responses, including the incorporation of mental health education into optometry curricula, the provision of accessible and confidential counselling services, and the development of targeted support strategies for high-risk groups. National regulatory bodies, such as the Optometrists and Dispensing Opticians Registration Board of Nigeria (ODORBN), should consider developing standardized mental health guidelines for training institutions. Additionally, inter-professional collaboration, improved infrastructure, and policy interventions addressing systemic contributors to student distress, such as academic instability and resource limitations, are essential. Public health authorities should prioritize student mental health in national strategies and implement evidence-based awareness and stigma-reduction campaigns. Finally, sustainable funding and support for longitudinal research are necessary to monitor trends, evaluate interventions, and inform policy and practice across tertiary institutions.

## Data Availability

All relevant data are within the manuscript and its Supporting Information files.

